# Refocusing Algorithmic Fairness on Feature-Level Bias: A Diagnostic Approach Using Dutch EHR Data

**DOI:** 10.1101/2025.11.09.25339863

**Authors:** Cathleen S. Parsons, Shelley-Ann M. Girwar, Sepinoud Azimi, Marco R. Spruit, Marcel R. Haas

## Abstract

While algorithmic fairness research in healthcare has predominantly focused on disparities in model performance, less attention has been given to the underlying data structures that may drive such disparities. High-level fairness metrics often obscure the deeper feature-level dynamics necessary for a critical and context-aware assessment of fairness. To address this gap, we propose and apply a diagnostic framework termed Feature-Level Bias Identification And Sensemaking (FL-BIAS). As a case study, we conducted a secondary analysis of a retrospective cross-sectional cohort study using electronic health record (EHR) data from Dutch general practitioners, linked with sociodemographic data from Statistics Netherlands. The dataset included 112,872 patients, of whom 16.2% had a non-Western migration background. Hospitalization in the following year was modeled using Johns Hopkins Aggregated Diagnosis Groups (ADGs). We trained logistic regression and XGBoost models on different subgroup datasets to evaluate performance disparities using fairness metrics. To analyze feature-level contributions to predictions and errors, we applied Shapley Value methods, including Kernel SHAP and Cohort Shapley. Exploratory analysis revealed significant differences in SES and ADG distributions between Dutch and non-Western groups, though Multiple Correspondence Analysis showed minimal structural variation. Mediation analysis indicated that the effect of migration background on hospitalization was largely mediated by SES, with potential unobserved confounding. While standard fairness metrics indicated modest bias in favor of non-Western patients, deeper feature-level analyses revealed subgroup-specific patterns of variable importance that suggest potentially less favorable underlying conditions. For instance, malignancy (ADG 32) had a stronger predictive impact among non-Western patients but contributed less to false negatives compared to Dutch patients, possibly reflecting structural disparities in cancer diagnosis and care. These findings highlight the need for contextual, multi-level evaluations of algorithmic bias. Fairness in healthcare AI must be approached as a socio-technical challenge, requiring multidisciplinary collaboration to uncover root causes and guide effective mitigation strategies.

**Author Summary:** Unfair algorithms often originate in the data itself. To help detect such hidden biases, we combined known data science methods into a diagnostic approach named Feature-Level Bias Identification And Sensemaking (FL-BIAS). Using Dutch general practitioner records linked with national demographic information, we explored how patient migration background corrected for socioeconomic status affect predictions of hospital admissions. We discovered that certain medical conditions, such as cancer and chronic illness, contributed differently to predictions for Dutch compared to non-Western patients, revealing subtle but important patterns in how data reflects social inequalities. Our goal was to move beyond simple fairness scores and better understand how inequalities can be hidden in health data, supporting the design of fairer and more transparent healthcare AI tools that truly serve all patients.

## Introduction

The field of algorithmic fairness remains conceptually unsettled. Despite ongoing debates over how fairness should be defined, which metrics should be prioritized, and which mitigation strategies are most effective (1–3), there is widespread recognition that deploying inadequately evaluated algorithmic models introduces serious risks, particularly in high-stakes domains such as healthcare (4, 5). Predictive models that perform differently across subpopulations risk reinforcing or exacerbating existing societal disparities, a phenomenon termed algorithmic bias(6). In response, numerous guidelines and ethical frameworks have emerged that position fairness as a central design principle, including the World Health Organization’s *Ethics and Governance of Artificial Intelligence for Health* (7), the European Union’s AI Act (8), and other national-level initiatives.

Over the past decade, substantial research on algorithmic fairness in healthcare has primarily focused on the definitional complexities of fairness and on developing technical bias mitigation strategies such as reweighting, resampling, fairness-aware training, threshold adjustment and calibration techniques (2, 3, 9). However, as Corbett-Davies et al. have argued (10), formal fairness definitions often fail to address structural disparities when applied without a nuanced, contextual understanding, potentially leading to decision policies that result in adverse outcomes. Therefore, fairness approaches need to be explicitly outcome-oriented and context-aware (10). This is particularly crucial in health applications, where sensitive attributes may serve as proxies for socioeconomic status (SES) (11, 12). For example, one study found that Black race was not associated with higher in-hospital mortality compared to White race, after adjusting for socio-demographic and patient health profiles upon admission (13). Yet such context dependent relationships are frequently omitted from fairness analyses, where mitigation strategies often result in suboptimal solutions (3, 14).

Another complication concerns the effectiveness of bias mitigation strategies, which depends heavily on the specific dataset and model context (11, 15, 16). A well-grounded understanding of relationships between features within the dataset can even help to overcome the trade-off between fairness and accuracy when applying mitigation strategies that modify the data. Leininger et al. demonstrate that causal pre-processing methods, which aim to approximate a counterfactual “fair world” where sensitive features are causally independent of the outcome, can improve fairness without negatively impacting accuracy by addressing bias at its structural source (17).

Both the dependency of fairness definitions on data context and the reliance of mitigation strategies on dataset structure highlight the need to understand where and how bias manifests in the data. Fairness methodologies are thus inherently vulnerable to the validity of assumptions made by human experts, including beliefs about relationships between features and outcomes, as well as the broader context in which the data was generated, such as the presence of proxy variables. To support domain experts in identifying patterns that require interpretation, and to inform the development of appropriate bias mitigation strategies, a systematic examination of feature behavior across subpopulations is essential, yet this step seems often overlooked.

Prior fairness assessments of electronic health record (EHR) data have largely focused on applying group-level fairness metrics rather than uncovering potential sources of bias at the feature level (2, 18). Yet complementing methods such as Shapley values, commonly used in explainable AI, can help reveal differences in feature responsiveness to prediction risks across subgroups in fairness analyses (19–21). Therefore, the objective of this study is to propose and apply a diagnostic approach to assess feature-level bias in predictive models trained on EHR data. A systematic examination of feature behavior offers a path beyond fairness metrics toward a human-in-the-loop assessment of fairness.

As a case study, we compared model performance and variable behavior between individuals from Dutch and non-Western migration backgrounds, controlling for SES, in Dutch primary care EHR data. A recent survey revealed that some Dutch patients with non-Western migration backgrounds perceive their health concerns as inadequately addressed by healthcare providers, potentially due to ethnic bias (22). This finding motivated the examination of whether such disparities are reflected in Dutch EHR data when developing predictive models. Through this case study, we successfully identified subgroup-specific differences in feature behavior that were not captured by standard fairness metrics.

## Methods

### Diagnostic approach

Figure 1 illustrates the proposed diagnostic approach Feature-Level Bias Identification and Sensemaking (FL-BIAS), which systematically assesses underlying data structures and feature behavior between subgroups. Interpreting data differences as indicators of algorithmic bias requires, as in causal inference, both data patterns and contextual understanding. Starting with a general understanding of the dataset’s structure, successive steps progressively uncover the features behind algorithmic bias.

**Figure 1:**
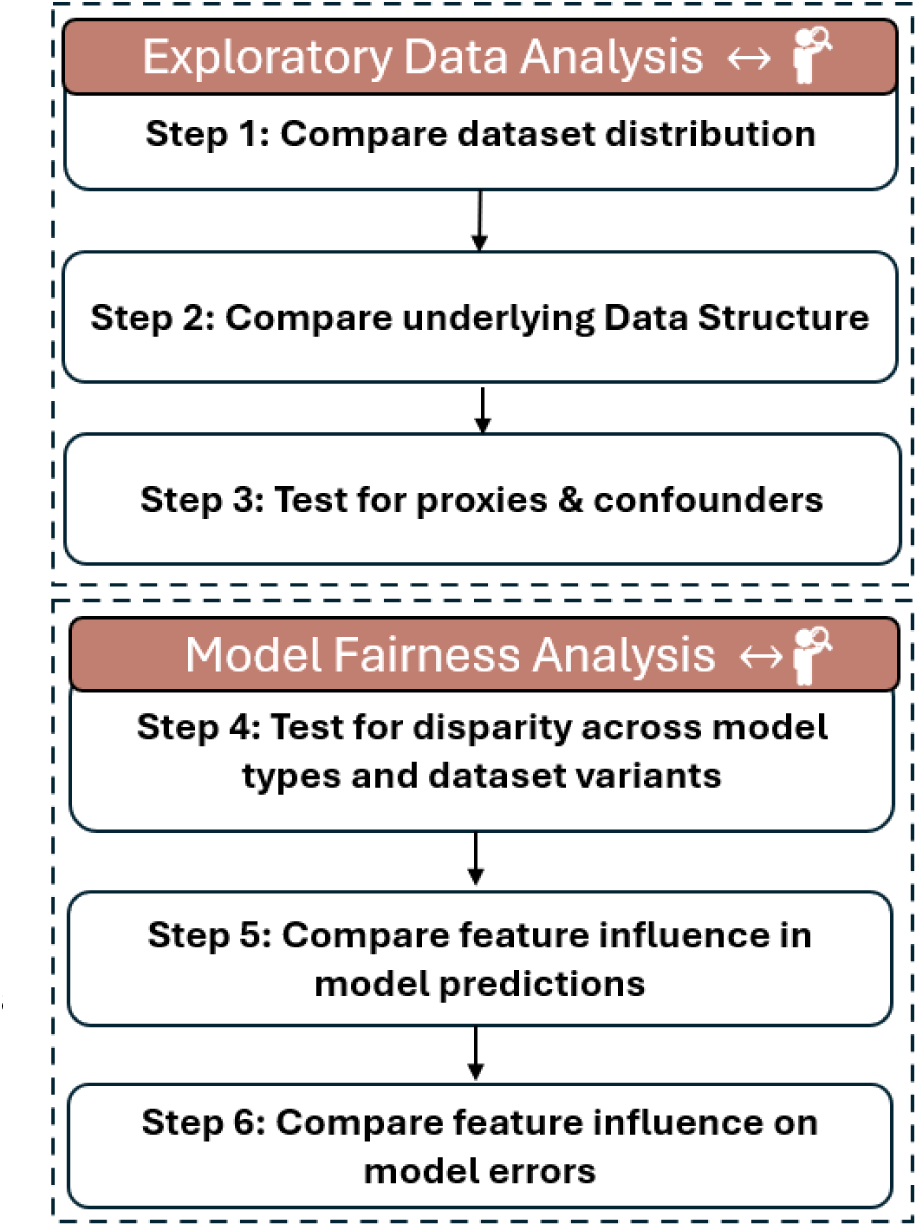
FL-BIAS (Feature-Level Bias Identification And Sensemaking): a diagnostic framework that systematically assesses underlying data structure and feature behavior between subgroups

### Exploratory Data Analysis Phase

The first phase provides a systematic examination of the datasets for the two subgroups.

Step 1: Evaluate subgroup distributions of demographic and clinical features to identify imbalances that may explain observed differences in model performance. This can include feature prevalence analysis, statistical tests, and visual inspections.

Step 2: Explore how features contribute to the overall data structure and how they co-occur, revealing relationships that could influence predictions. This may involve Multiple Correspondence Analysis (MCA) or Principal Component Analysis (PCA).

Step 3: Account for potential confounders to correctly interpret the effect of sensitive attributes on outcomes and to determine which variables should be included in prediction models. Possible approaches include Causal Mediation Analysis or constructing data-driven Directed Acyclic Graphs (DAGs).

### Model Fairness Analysis Phase

The second phase focuses on model evaluation and identifying the extent of model performance bias at the feature level.

Step 4: Develop and evaluate models across multiple subdatasets and use diverse performance metrics to detect subgroup-specific disparities. Compare metrics such as Equalized Odds, False Negative/Positive rates, calibration curves, and ROC-AUC across groups to identify performance gaps.

Step 5: Assess relative feature importance differences between subgroups to identify uneven model dependencies. For instance, by comparing coefficients or permutation-based feature importances, or by applying XAI techniques such as Kernel Shapley Values to quantify how features contribute differently to predictions across groups.

Step 6: Examine model prediction errors across groups to identify which features drive observed disparities. This can involve analyzing misclassification patterns by feature values or using error attribution analysis such as Cohort Shapley Values.

Collectively, these steps highlight differences in data that may reflect inequities between subgroups. Recognizing such inequalities, however, requires human interpretation informed by clinical and social context. The following case study describes how this diagnostic framework was applied to real-world data, outlining the methods used to operationalize each above mentioned step.

### Case Study Design

This study was a secondary analysis of a retrospective cohort study (23, 24). The EHR data from Dutch general practices, covering the period from January 2016 to December 2016, were linked in a controlled environment with data from Statistics Netherlands, the Dutch central bureau for statistics. These included 2016 data on migration background and SES, as well as 2017 data on hospitalization outcomes.

### Study Population

General practices across four diverse and representative regions in the Netherlands provided data from their patient EHRs containing ICPC-1 diagnosis codes (Dutch version of International Classification of Primary Care) and medication prescriptions (Anatomical Therapeutic Chemical codes, ATC-4). To reduce data dimensionality, the Johns Hopkins Adjusted Clinical Groups (ACG) system, version 12.1, was used to categorize ICPC-1 diagnosis codes into 32 Aggregated Diagnosis Groups (ADGs) based on comparable burden of care, which is translated from amongst others complexity and intensities of health conditions (25). As prescribing behavior, typically guided by well-established treatment protocols, may diverge from diagnostic entries in electronic health records, the ACG system was additionally used to generate 81 Rx-defined Morbidity Groups (Rx-MGs) from prescription data. This dual approach has been used in previous studies (26, 27), and is applied here to identify distinct sources of potential bias in EHR data by comparing models developed from diagnostic data with those based on prescribing data.

Our EHR data were linked with three datasets from Statistics Netherlands: 2017 hospitalization data and 2016 data on migration background and SES. For migration background, a dataset was used that assigns each subject in the Netherlands to either their own country of birth or, if born in the Netherlands, to the country of birth of their non-Dutch parent (with the mother’s country used by default if both parents are non-Dutch). This database has been used in a previous Dutch study researching the relation between ethnicity and population cardiovascular health (28). For our own analysis, we divided the database, which contained no missing values, into two groups: Dutch and non-Western country codes with at least 500 people per country. Patients with a Western migration background were excluded to maintain a clear analytical contrast between the study groups that formed the focus of the fairness analysis.

For SES, similar to the approach used in a previous Dutch population-based study (28), we used data based on the percentile ranking of household prosperity, which included standardized disposable household income and wealth. CBS data on occupation and education were not included in the analysis of SES, as they contained a high degree of missing data (30%). Following that previously published approach (28), we categorized the lowest 20% as low SES, the highest 20% as high SES, and the middle 60% as middle SES, using the distribution in the total dataset (including participants with Western backgrounds). The SES variable had only 0.6% missing data, which were imputed using the mode.

Due to the imbalanced nature of subgroups of the EHR dataset, a subset with an equal distribution of Dutch and non-Western populations was created as an additional dataset for comparative analyses. Additionally, given that many patients are relatively healthy and thus have low rates of hospitalisation, a subset of high-morbidity patients (i.e., those with at least one major ADG diagnosis) was constructed for subgroup-level analysis.

### Analysis

As outlined in Figure 1, we first compared the prevalence distributions of ADGs between Dutch and non-Western populations. Second, to assess the underlying data structure, we performed a Multiple Correspondence Analysis (MCA) using the ‘FactoMineR’ and ‘factoextra’ R packages. Both sub-datasets had an equal distribution of the confounders age, sex, SES and hospitalization outcomes . MCA reduces categorical data to latent dimensions that capture variance in feature associations, and visualizes how categories cluster, thereby revealing potential co-occurrence patterns. Squared cosine (cos^2^) values are added, which measures the degree of association and indicate how well each feature is represented by the first two dimensions. Third, we conducted a mediation analysis with the ‘Mediation’ R package (29) to examine the extent to which the effect of migration background on hospitalization was mediated by SES. Sex and age are potential confounders influencing

SES and hospitalization risk and included as co-variates. Clinical risk factors, in our context ADGs, were not included, as they are considered to lie on the causal pathway from migration background to hospitalization, potentially operating through SES, and including them would block part of the mediating effect. Causal mediation analysis relies on the assumption of sequential ignorability (29), which requires the absence of unmeasured confounders and necessitates sensitivity analysis. We choose as the sensitivity parameter (ρ) for unobserved confounding the correlation between the residuals of the mediator and outcome regressions (29). The sensitivity analysis was conducted by varying the value of ρ in increments of 0.1.

For the Model Fairness Analysis Phase of our framework (Figure 1), we developed multiple models as outlined in Figure 2. To control for potential confounding effects, age, gender and SES were incorporated as variables in our models. We selected hospitalization (prevalence of 10% in our dataset) in the following year, one of the standard predictions by the ACG system, as the outcome variable. For interpretability of feature importance and based on (23)’s approach using logistic regression with ADGs, we selected logistic regression as our baseline model for predicting hospitalization. To account for any non-linear relationships and interactions between the features, we compared the logistic regression model to a XGBoost model. Additionally, a logistic regression model utilizing only eight major ADGs (3, 4, 9, 11, 16, 22, 25, 32) was developed. Major ADG models are commonly employed in practice and offer the advantage of reducing data dimensionality and improving explainability. Model specifications are detailed in S1 Table. During development, no threshold adjustments were applied, as the primary aim was not to optimize predictive performance for imbalanced datasets but to examine differences in model behavior between the two subgroups using identical model architecture.

**Figure 2:**
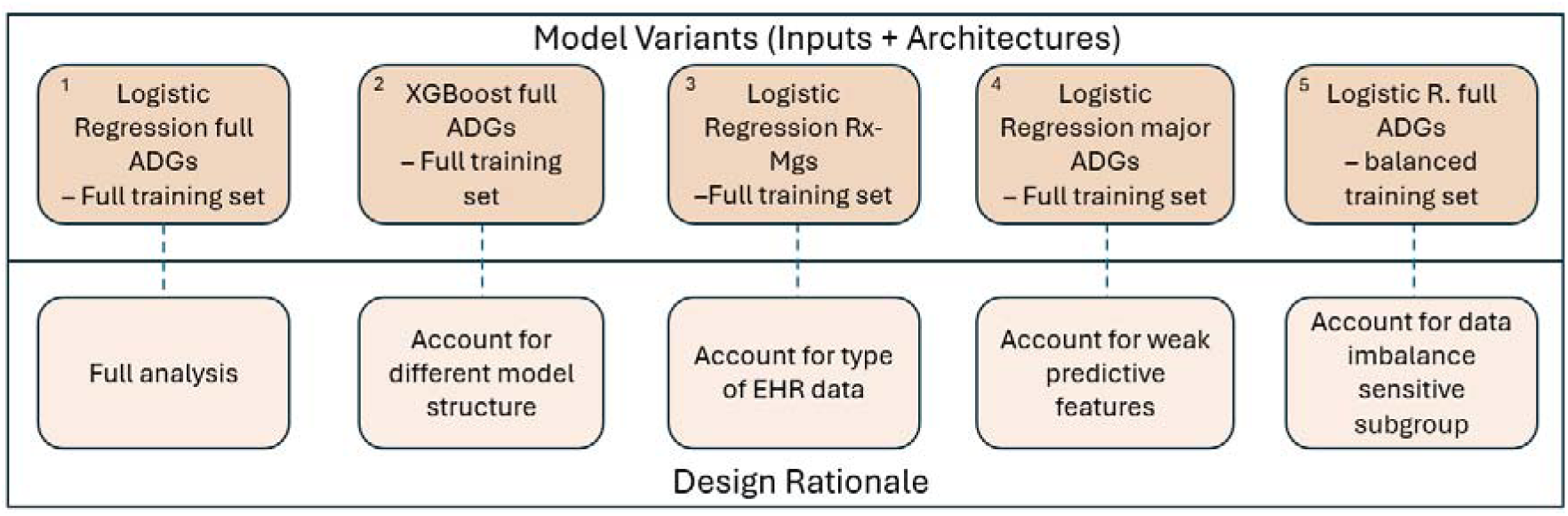
overview model variants (Inputs + Architectures) and its rationale

The models of Figure 2 were assessed on several performance metrics (described in the following section). In addition, the LR–32 ADGs, LR–balanced dataset, and RX-MG models were evaluated on Dutch-only and non-Western-only datasets. Feature importance differences were examined specifically for the LR–32 ADGs model, both on the full dataset and in subgroup-specific models for Dutch and non-Western patients. To ensure reliable coefficient interpretation in these logistic regression models, multicollinearity analyses were performed using the Variance Inflation Factor (VIF) and correlation heatmaps. Lastly, we conducted analyses using Shapley Values (SV). SV are being increasingly used in the algorithmic fairness field to quantify contributions of individual variables to algorithmic bias (19, 30). For this analysis, only major ADGs were selected to limit computational expense. We calculated two types of Shapley Values: Kernel Shapley Values, implemented using the ’Shapr’ package in R (31), and Cohort Shapley Values, implemented using the ’cohort shapley’ Python package(30). By employing computationally efficient Kernel Shapley Values calculations from ‘Shapr’ (31), we were able to compare the responsiveness of each variable to the model’s prediction between the Dutch and non-Western datasets. Kernel Shapley Values estimate the contribution of each feature by averaging its marginal effect across all possible permutations of feature inclusion. Cohort Shapley was used to measure SV to instances where the model predicted no event but one actually occurred (false negatives) and for the differences between observed and predicted outcomes (residuals). This method by (21, 30) uses only observed combinations in data, thereby avoiding unrealistic feature combinations, and accounts for both feature interactions and dependencies.

### Outcome

Fairness can be defined in multiple ways that are theoretically incompatible when outcome prevalence differs across groups (32). Overall, a distinction is drawn between whether the desired outcome involves equal performance or distributive justice, which refers to equal allocation. In the setting of this study, it is preferred that the model performs comparably across all groups, which is generally used for predictive models in the healthcare field. For predicting hospitalisation, we are particularly interested in false negatives rates (FNR) as metric, since it can be used to measure undertreatment in the minority groups (33). Equalized odds (EO) (34) is a commonly used fairness metric, ensuring that true positive rates and false positive rates are balanced among subgroups. An EO ratio of 1 indicates complete fairness. EO was calculated via the Fairlearn package (35). Fairness metrics were computed on the complete test set using the Fairlearn toolkit, following their recommended approach for subgroup analysis.

To evaluate 5-fold cross-validation model performance on an imbalanced dataset, we included several additional metrics beyond FNR, accuracy and the C-statistic. These included Precision (TP / [TP + FP]) and the F1-score (2 × Precision × Recall / [Precision + Recall]), which balances the trade-off between Precision and Recall (TP / [TP + FN]). The Matthews Correlation Coefficient (MCC) was also reported, as it is particularly well-suited for imbalanced data. MCC considers all four quadrants of the confusion matrix, True Positives (TP), True Negatives (TN), False Positives (FP), and False Negatives (FN), proportionally considering both the positive and negative elements within the dataset. It is defined as: ((TP×TN-FP×FN)/ √((TP+FP)×(TP+FN)×(TN+FP)×(TN+FN))). The MCC ranges from –1 to +1, where +1 indicates perfect prediction, 0 indicates performance equivalent to random chance, and –1 indicates complete disagreement between predictions and observations.

### Ethical considerations

This analysis used the general practice data from a previous project (23, 24). For that project, The Medical Ethics Committee of Leiden University Medical Center (reference G19.048) determined that formal ethical review was not required, as it involved retrospectively and routinely collected, de-identified health data.

Processing special-category data such as health and migration background was carried out in accordance with GDPR Article 9(2)(j), which permits such processing for scientific research in the public interest when appropriate safeguards are in place. All analyses were performed exclusively within the secured Statistics Netherlands secured environment, and researchers had no access to directly identifiable data at any time. Patients who could not be linked to the Statistics Netherlands database and deceased patients, who could not exercise the right to object to data use, were excluded. The output of all analyses was subject to Statistics Netherlands disclosure control to ensure that all published results were sufficiently anonymized and posed no risk of re-identification.

## Results

### Step 1: Compare dataset distribution

The total study population of our case study consisted of 112,872 patients of which 16% were identified as non-Western and 84% as Dutch. A significant difference was observed in the SES of non-Western individuals, marked by a 20% prevalence percentage point difference to Dutch individuals. In terms of morbidity, the Dutch population, which was on average older, exhibited a higher prevalence of ADGs and a greater frequency of hospitalization.

S2 Table provides an overview on the prevalence distribution of ADGs per population dataset. Comparing the prevalence in both datasets, ADG1, ADG 5, and ADG 7 had the largest difference, with more than 5% percentage prevalence point difference and being statistically significant.

### Step 2: Compare underlying data structure

Figure 3A and 3B visually represent the contribution values for each ADG’s presence and absence across the first two dimensions and their clustering. The plots represent a modest 15% of the dataset variance, consistent with high-dimensional categorical clinical data where variance is distributed across many components. The primary dimensions for both the Dutch and the non-Western dataset presented comparable but relatively low eigenvalues, indicating that no single combination of ADGs accounts for a large portion of the variability within the data (S3 File). Features with low cos² values, such as ADG +17 and ADG +18, are poorly represented by the first two dimensions. Therefore, any apparent shift in their positions between the Dutch and non-Western plots of Figure 3 should not be interpreted as meaningful. However, ADG +14 shows a notable positional shift with a modest cos² value, suggesting different co-occurrence patterns between groups.

**Figure 3.**
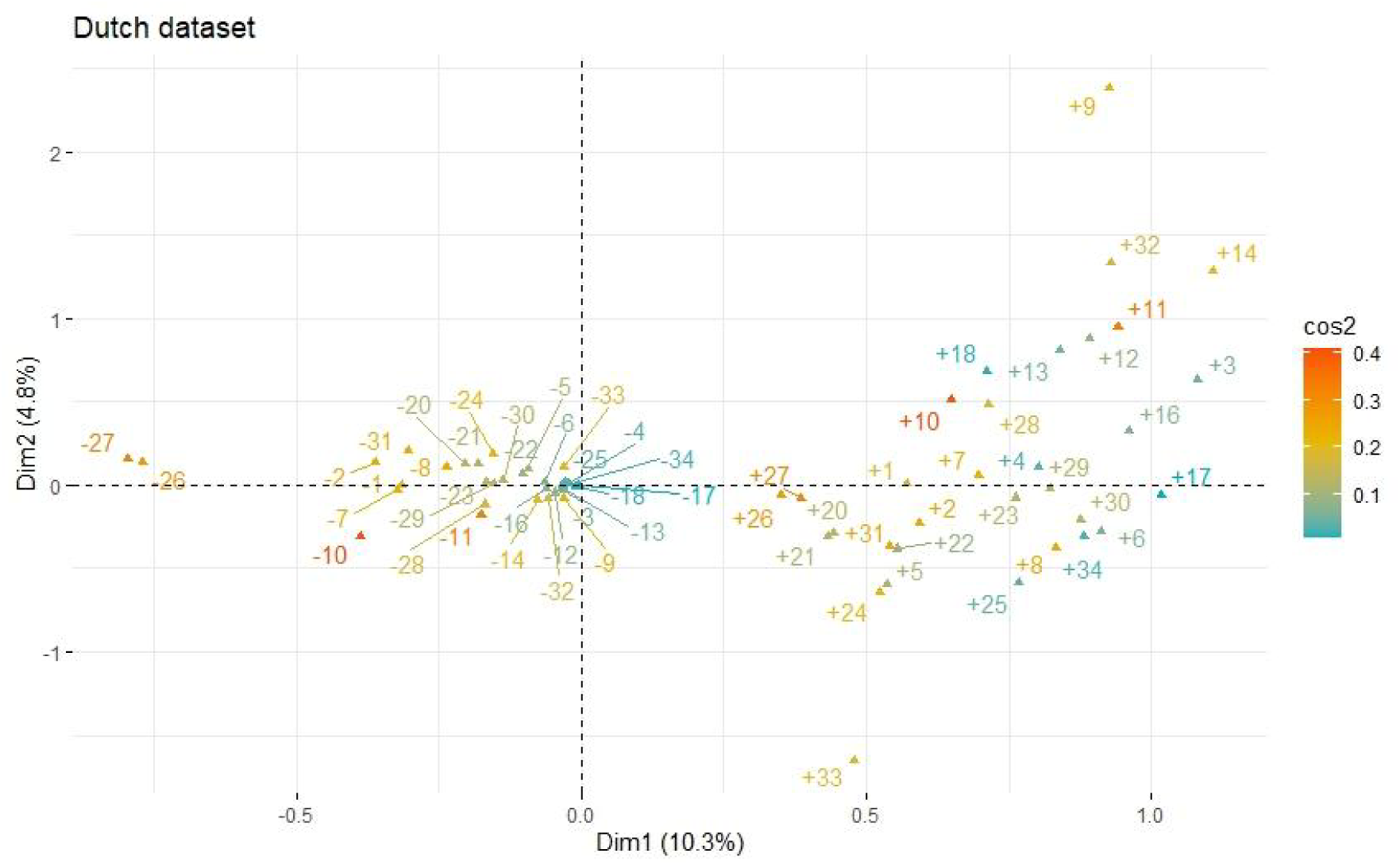
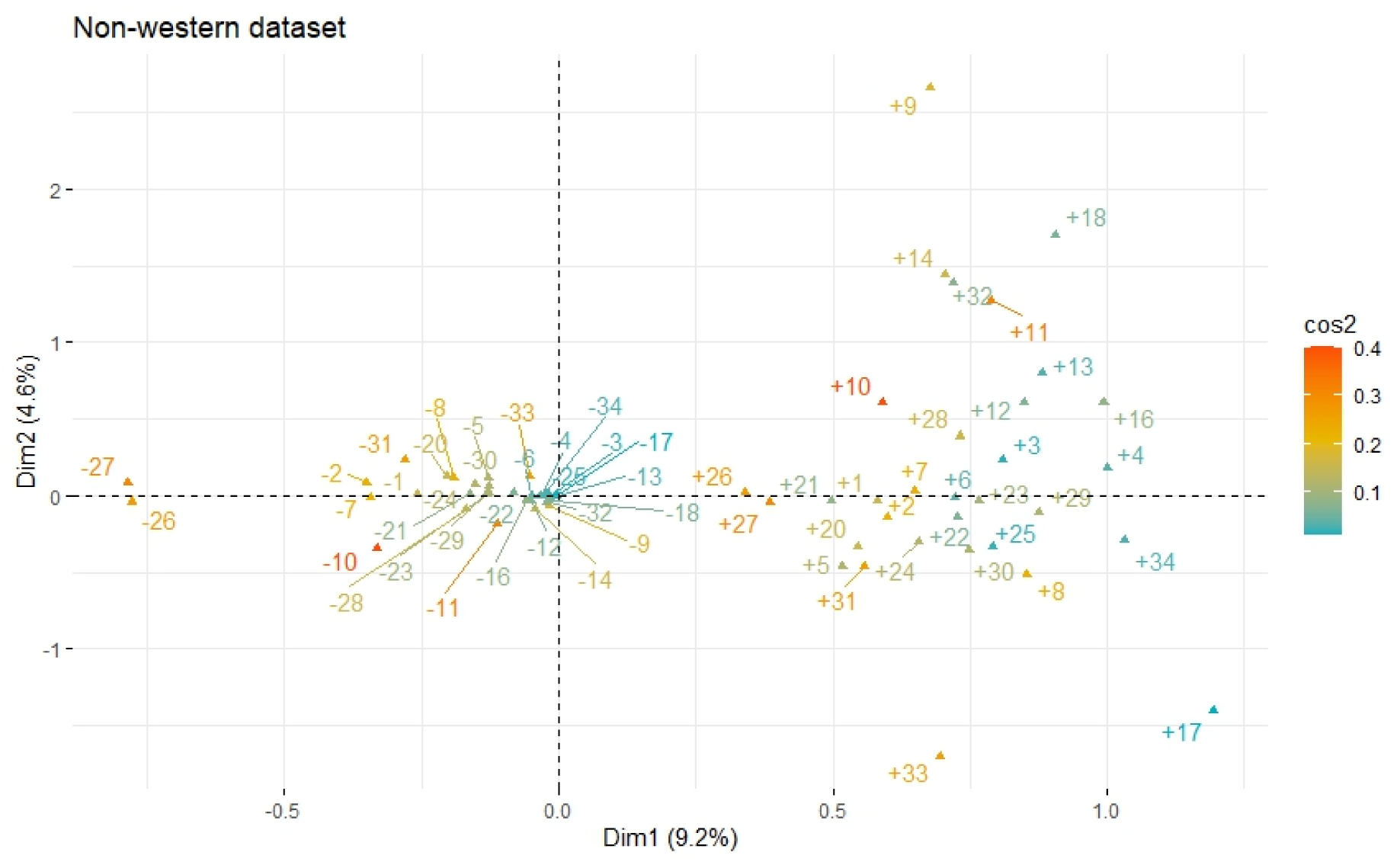
Multiple Correspondence Analysis of ADG variables in Dutch dataset (3A) and non-Western dataset (3B). Triangles indicate feature positions (co-occurrences), with “+” for ADG present and “-” for ADG absent. Cos^2^ values measures the degree of association and indicate how well each feature is represented by the first two dimensions.

### Step 3: Test for proxies & confounders

The Average Causal Mediated Effect (ACME), estimating whether non-Western background is associated with higher hospitalization rates through lower SES, was statistically significant for both the treated non-Western group (0.005695) and the control Dutch group (0.005758), with p-values less than 0.001. In contrast, the Average Direct Effect (ADE), which captures the direct effect of migration background on hospitalization after accounting for SES, was not statistically significant (p = 0.68). The proportion mediated exceeded 1.0 (≈1.35), consistent with full mediation and suggestive of a suppression effect, although this pattern is based on a non-significant direct effect. In sensitivity analysis, the mediation effect (ACME) was eliminated when ρ reached −0.1, revealing that a small degree of unobserved confounding could nullify the mediation effect. Additional estimation details are provided in S4 Table.

Overall, the mediation analysis indicates that migration background does not directly affect hospitalization. Instead, the statistically significant but small disparities appear to be fully explained by socioeconomic differences, with possible contributions from other unmeasured factors.

### Step 4: Test for disparity across model types and dataset variants

An overview of the performance metrics for the regression models can be found in Table 2. Despite correction for imbalanced data during model training, the performance of each model, particularly the XGBoost, was found to be suboptimal. This is primarily demonstrated by the low precision scores. Given that 90% of the patients were not hospitalized and major ADGs are indicative of hospitalization, further analysis was conducted on a sub-dataset consisting exclusively of patients with high morbidity, identified by the presence of one or more major ADGs. While this approach enhanced precision, it also led to a slightly higher algorithmic bias for both the full model and the major ADG model. This increase in disparity also occurred when reducing the model ADG variables to the eight major ADGs.

For the models based on ADG variables, the EO ratio was below one and non-Western individuals had a slightly higher FNR. This disparity effect remained when the models were trained on the balanced dataset. Overall, the logistic regression ADG-32 model demonstrated slightly better performance metrics for the non-Western population (Table 2), as reflected in a Matthews parity value of 1.051 (S5 Table) and illustrated by ROC curve analyses for each subgroup (Figure 4). Conversely, the logistic model trained on medication prescription data (Rx-MG) exhibited a lower degree of disparities and demonstrated a slightly more favorable performance when applied to the Dutch population (Table 2, Figure 4).

**Figure 4:**
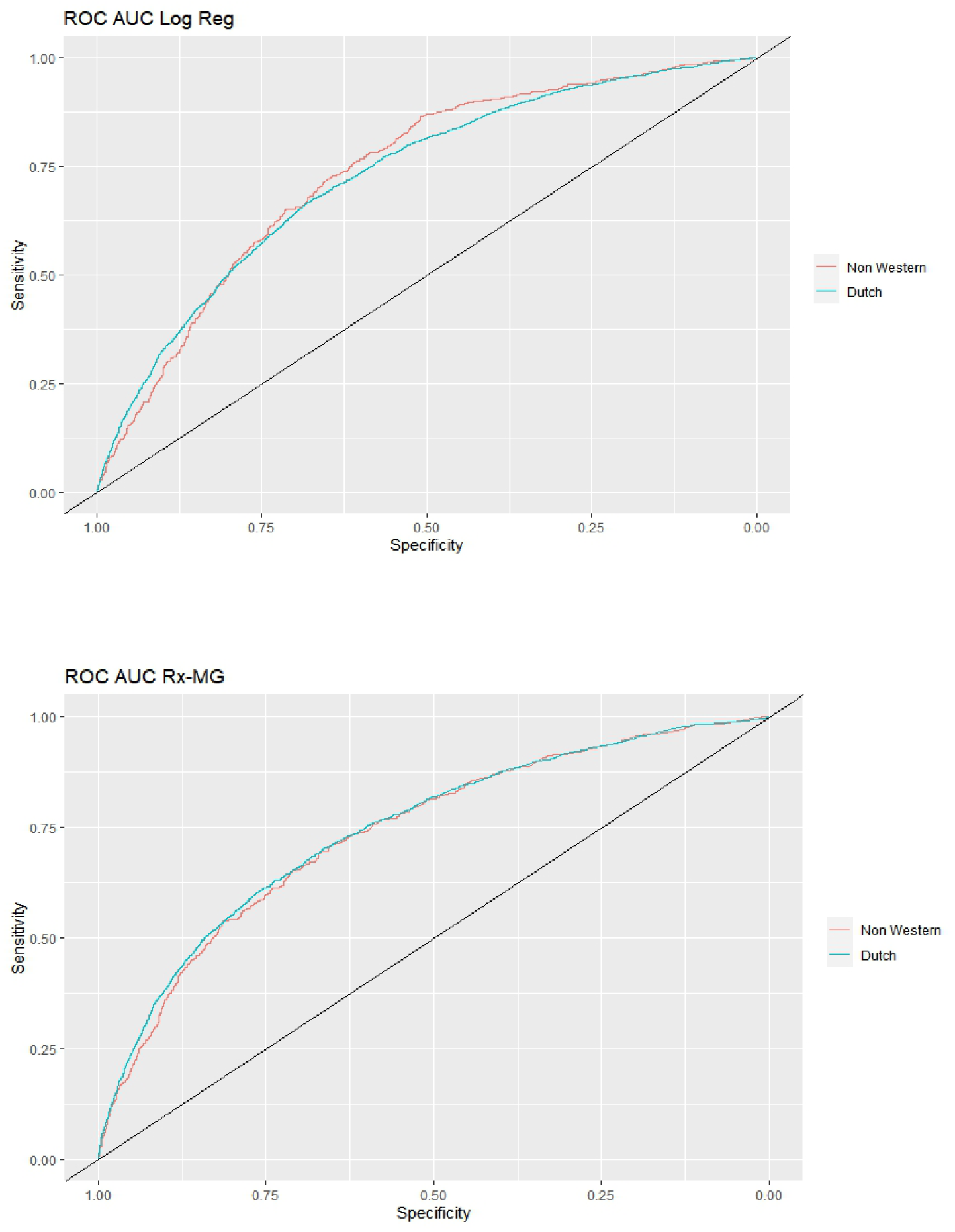
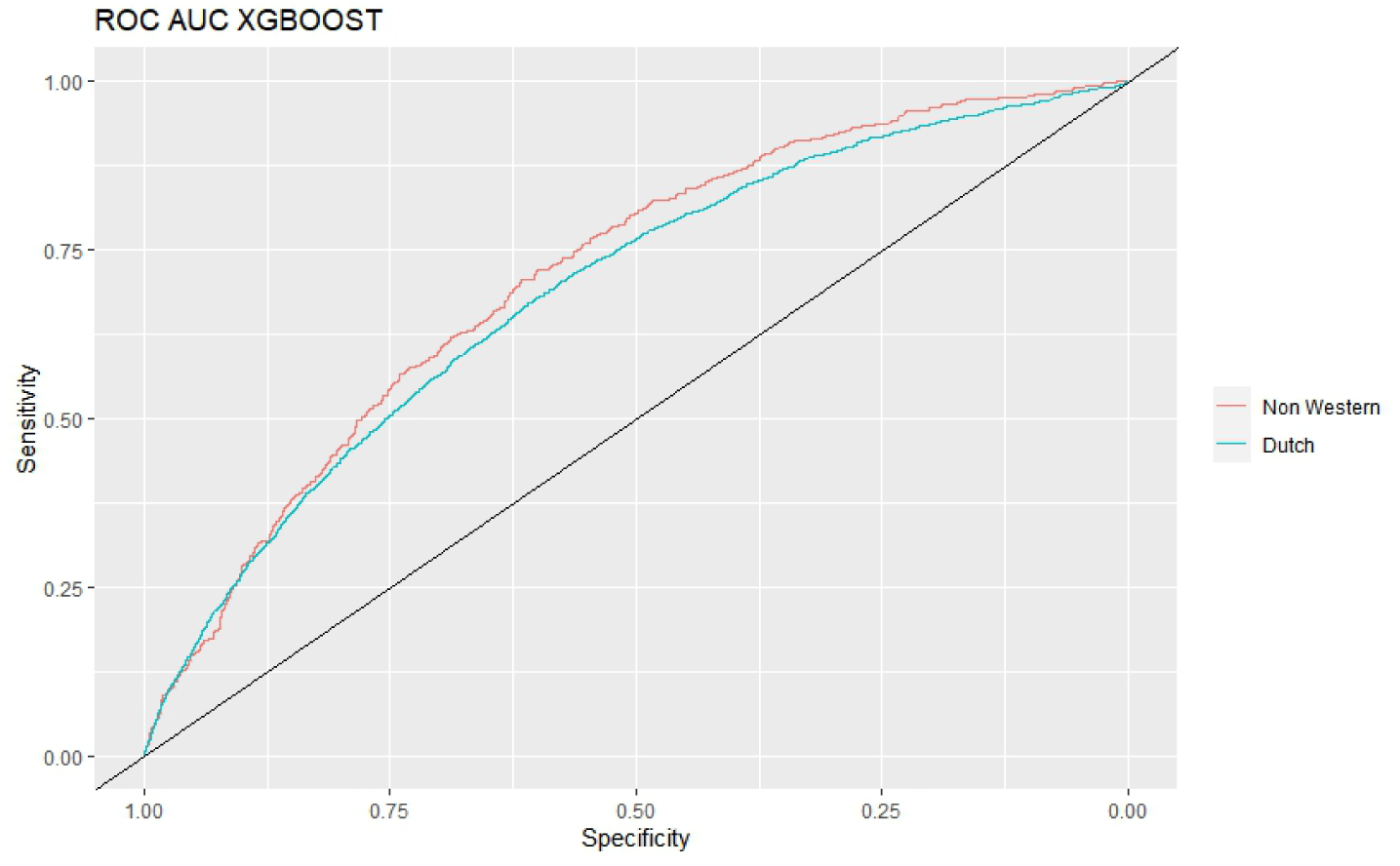
ROC-curve Logistic Regression 32 ADG (4A): ROC AUC-curve logistic regression Rx-MG(4B); ROC-curve XGBoost ADG (4C)

### Step 5: Compare feature influence and responsiveness in model predictions

The VIF analysis showed minimal multicollinearity among ADG features, with a mean VIF of 1,094 (SD = 0,075) (S6 Table). This was supported by the low pairwise correlations between ADGs (S7 Figure), where the highest observed correlations were between ADG 10 & 11, ADG 26 & 27 and ADG 7 & ADG 11. Logistic regression coefficients (S6 Table) identified ADG 33 (Pregnancy), ADG 32 (Malignancy), ADG 11 (Chronic Medical: Unstable), and ADG 9 (Likely to Recur: Progressive) as the strongest predictors of hospitalization. With the exception of pregnancy, these ADGs belong to the Major ADG category. Kernel Shapley Value analysis (Figure 5; S8 Table) of the major ADG model further highlighted the importance of ADG 11, ADG 32, and ADG 9 in the model’s predictions.

**Figure 5:**
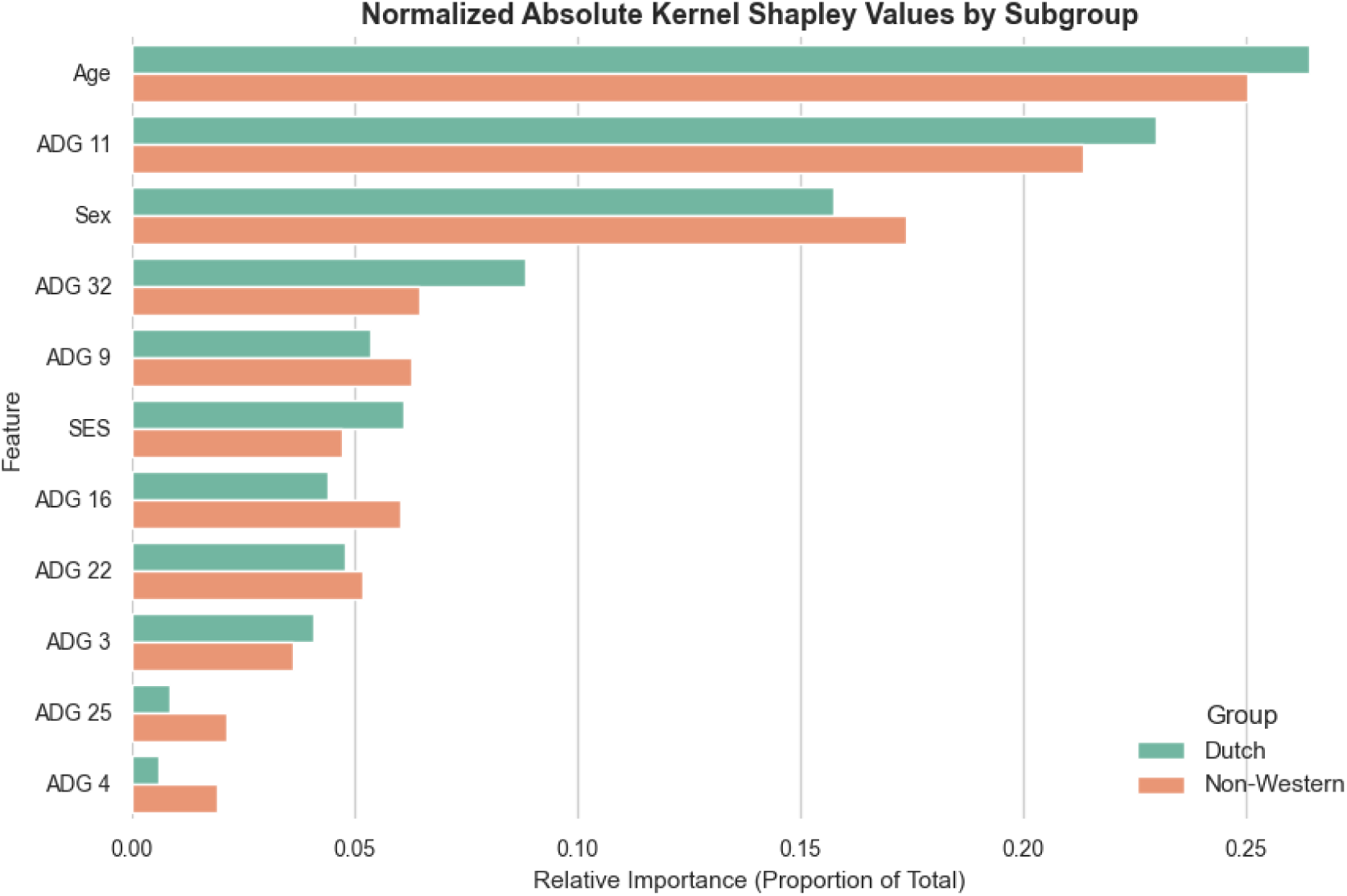
Kernel Shapley Values; Subset of data used, with 1000 positive event (hospitalization) and 1000 negative event (no hospitalization)

In the major ADG model trained on the Dutch dataset, SV for both ADG 32 and ADG 11 were higher than in the model trained on the non-Western dataset. For ADG 32, the Dutch model showed higher absolute sum of Shapley Values (57.3 vs. 37.6) even though the logistic regression coefficient was lower in the Dutch full ADG model (0.510 vs. 0.742), reflecting its greater prevalence in the Dutch dataset (5.6% vs. 2.6%; Table 1). Similarly, ADG 11 was more prevalent among Dutch patients (14.6% vs. 11.6%; Table 1), which corresponds to higher Shapley Values in the Dutch major ADG model (148.9 vs. 124.6) despite comparable coefficients across groups (0.520 vs. 0.515).

**Table 1:** patient characteristics in the retrospective dataset

|  | Overall | Dutch | Non-Western |
| --- | --- | --- | --- |
| n (%) | 112872 (100) | 94628 (83.8) | 18244 (16.2) |
| Age (mean (SD)) | 45.92 (19.58) | 46.54 (19.78) | 42.74 (18.20) |
| Male (%) | 55356 (49.0) | 46694 (49.3) | 8662 (47.5) |
| SES (%) |  |  |  |
| *High* | 27627 (24.5) | 25400 (26.8) | 2227 (12.2) |
| *Middle* | 71793 (63.6) | 61093 (64.6) | 10700 (58.6) |
| *Low* | 12787 (11.3) | 7596 ( 8.0) | 5191 (28.5) |
| *NA* | 665 ( 0.6) | 539 ( 0.6) | 126 ( 0.7) |
| GP visits (mean (SD)) | 3.43 (4.26) | 3.40 (4.27) | 3.62 (4.21) |
| Hospitalized (%) | 10955 ( 9.7) | 9290 ( 9.8) | 1665 ( 9.1) |

**Table 2:** Performance metrics. Model column indicates type of model, where LR stands for Logistic regression. Models were trained on the trainingsdataset of 84.654 patients of which 16% non-western, if not specified otherwise. The balanced dataset consisted of 13592 Dutch and 13592 non-western patients. EO: equal odds ratio. FNR: False Negative Rate** Standard deviations across standard metrics were all < 0.02 and are not displayed for clarity. N.A.: Not Applicable

| Model | Test sample | F1 score | Precision | Accuracy | ROC-AUC | EO | FNR |
| --- | --- | --- | --- | --- | --- | --- | --- |
| LR - 32 ADGs | Full (28218) | 0.286 | 0.183 | 0.684 | 0.729 | 0.899 | 0.335 |
| LR - 32 ADGs | Dutch (23566) | 0.286 | 0.182 | 0.678 | See Fig. 4 | n.a. | 0.333 |
| LR - 32 ADGs | Non-western (4652) | 0.290 | 0.187 | 0.706 | See Fig. 4 | n.a. | 0.349 |
| LR - 32 ADGs | >= one major ADG (10492) | 0.341 | 0.233 | 0.623 | 0.675 | 0.878 | 0.366 |
| LR - major ADGs | Full (28218) | 0.268 | 0.173 | 0.684 | 0.693 | 0.820 | 0.421 |
| LR - major ADGs | >= one major ADG (10492) | 0.329 | 0.223 | 0.610 | 0.657 | 0.775 | 0.383 |
| LR - 32 ADGs - balanced dataset | Full (28218) | 0.285 | 0.183 | 0.689 | 0.729 | 0.904 | 0.329 |
| LR - 32 ADGs - balanced dataset | Dutch (23566) | 0.285 | 0.179 | 0.67 | 0.674 | n.a. | 0.326 |
| LR - 32 ADGs - balanced dataset | Non-western (4652) | 0.288 | 0.185 | 0.700 | 0.68 | n.a. | 0.344 |
| LR Rx-MG | Full (28218) | 0.306 | 0.204 | 0.734 | 0.680 | 0.971 | 0.387 |
| LR Rx-MG | Dutch (23566) | 0.309 | 0.206 | 0.734 | See Fig. 4 | n.a. | 0.384 |
| LR Rx-MG | Non-western (4652) | 0.294 | 0.195 | 0.734 | See Fig. 4 | n.a. | 0.402 |
| XGBoost - 32 ADGs | Full (28218) | 0.164 | 0.147 | 0.818 | 0.709 | 0.925 | 0.813 |
| XGBoost - 32 ADGs | Dutch (23566) | 0.167 | 0.150 | 0.815 | See Fig. 4 | n.a. | 0.808 |
| XGBoost - 32 ADGs | Non-western (4652) | 0.150 | 0.139 | 0.830 | See Fig. 4 | n.a. | 0.837 |

In contrast, in the non-Western dataset, ADG 4 (Time Limited: Major-Primary Infections), ADG9 (Likely to Recur: Progressive), ADG 16 (Chronic Specialty: Unstable Orthopedic) and ADG 25 (Psychosocial: Recurrent or Persistent, Unstable) had higher Shapley Values, indicating their stronger contribution to model predictions in this population.

### Step 6: Compare Error attribution features

The Cohort Shapley Values (Figure 6; S9 Table) specifically measured feature contributions to the prediction errors based in only observed combinations in data. Notably, ADG 11 appeared particularly influential in contributing to disparate impact within false negative cases, but played a less prominent role in overall residuals. Overall, the FNR tended to be higher in the non-Western group, aligning with higher FNR fairness metrics for that subgroup (Table 2), indicating under-recognition of hospitalization risk among non-Western patients.

**Figure 6:**
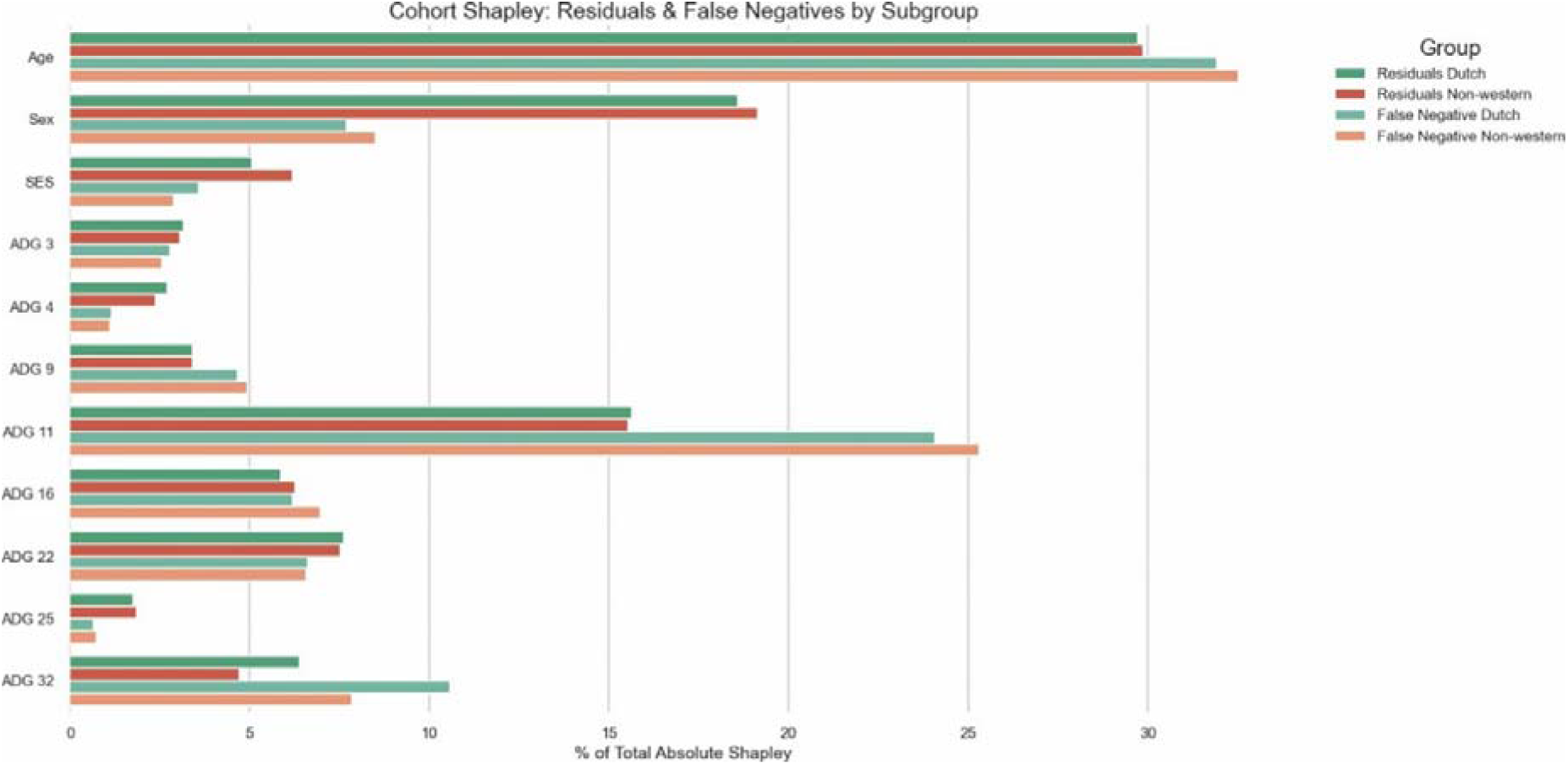
Cohort Shapley Values to calculate feature contributions to residuals, false negative cases and true label (actual hospitalization in 2017); Subset of dataset with only hospitalization outcome resulted into 2604 individuals. Balance was slightly skewed (1359 Dutch and 1235 Non-Western), so equal groups of 1235 were sampled.

As described in the previous step, ADG 32 was less prevalent in the non-Western population but had a stronger coefficient. The lower responsiveness of ADG 32 for residuals and false negatives specifically compared to in the Dutch population (Figure 6; S9 Table) further suggest that, when malignancy was diagnosed, it was more strongly associated with hospitalization outcomes in the non-Western group. This indicates subgroup-specific differences in how ADG 32 influenced model predictions.

## Discussion

Algorithms learn patterns in data. As a result, algorithmic fairness depends on awareness of the (societal) patterns reflected in dataset structures. While performance differences between subgroups may partly reflect variance in outcome prevalence, which is difficult to overcome, systematic bias in the data, such as differences in diagnostic documentation or treatment practices, is potentially correctable when detected. In our study, we systematically examined two subpopulations defined by migration background using our Feature-Level Bias Identification And Sensemaking (FL-BIAS) framework. We demonstrated that, beyond standard performance metrics, valuable insights can be obtained by integrating complementary data science methods. Particularly, we highlighted the added value of using Shapley analyses: Shapley Values can help uncover feature-level bias patterns that would otherwise remain hidden in aggregate metrics. While this particular case study relied on ADGs from the widely used Johns Hopkins ACG system, our fairness diagnostic approach FL-BIAS is broadly applicable to any other structured representations or clinically meaningful subsets of EHR data.

The main strength of our case study was the use of data from Statistics Netherlands, which enabled us to adjust for SES and to create subpopulations of different migration backgrounds. Our developed models using ADG classification appeared to have mild degrees of unfairness, which in this context is defined as a disparity in FNR and EO ratio. Discrepancies in FNR among groups implied that the model underestimates or encounters more difficulty in identifying positive outcomes for a subgroup. When predictive models were trained on pooled data (from Dutch and non-Western combined), the model learned a population-average association between features and outcomes. As a result, it tended to underestimate hospitalization risk for non-Western patients, contributing to higher FNR in this group. Unequal EO ratios entails an uneven distribution of true positives and false positives across different subgroups. The slightly lower EO ratio observed in our study (below one) may reflect several factors, including disparities in the distribution of ADG variables between groups, potential diagnostic differences, and variations in how these features relate to hospitalization outcomes.

While the ADG model performed slightly better overall performance in the non-Western population, a contrasting trend was observed for the Rx-MG model, which is based on medication prescription data. Reducing the ADG model to the eight major ADGs increased unfairness slightly. We further observed that certain ADGs were assigned different weights in the logistic regression model trained on the Dutch dataset as compared to the one trained on the non-Western dataset. The Shapley Values analysis highlighted a different pattern for ADG 32 (Malignancy) and ADG 11 (Chronic Medical: Unstable). While ADG 32 had higher coefficients in the non-Western model, it contributed less to model’s error compared to the Dutch database. This could be potentially explained by the well-documented disparities in screening participation for cancer (36), where non-participation can lead to more advanced forms of cancer in the non-Western population at time of diagnosis, consequently establishing stronger associations with hospitalization outcomes. Such a pattern may reflect a form of negative legacy bias (3), where delays in diagnosis become encoded in the data and influence subsequent algorithmic predictions.

It should be highlighted that the true extent of fairness in our case study remains unknown, since the sensitive variable, migration background, could have had a direct effect on hospitalization. This would have given a different threshold for hospitalization outcome per subpopulation and could have resulted in misrepresentation of FNR and EO, as discussed by Oneto (37). From our mediation analyses on our dataset, the direct link of migration background on hospitalization appeared not to be statistically significant. However, since variables such as health-care access were unobserved, hospitalization may not accurately represent a patient’s underlying health need. Regarding the ACME analysis, our SES did not include education levels due to high levels of missing data. This exclusion may have reduced the validity of SES as a proxy measure and contributed to the fragility of the ACME estimate, which was nullified at a rho of 0.1.

Our findings should therefore be viewed as an initial exploratory analysis of the extent of algorithmic bias in the real-world EHR data, serving as an example of how to begin uncovering the root causes of disparate outcomes. The main limitation was its models’ underperformance, which reduced the reliability and interpretive validity of the fairness metrics, feature importance estimates, and Shapley Value analyses. The relatively low F1 scores suggested that models based solely on ADG variables provide limited discriminative power for predicting hospitalization. In particular, our XGBoost model performed poorly due to the combination of many weakly informative ADG predictors, data sparsity, and class imbalance, as well as the poor probability calibration often observed in boosted trees (e.g.,38, 39), which together most likely led to diffuse decision boundaries. Deep learning architectures specifically designed for tabular data, such as TabNet (40), may have better handled the combination of sparse uninformative features and class imbalance. The ACG system, in practice, uses multiple sources of data for risk prediction, such as previous health care utilization and medication prescription in addition to the diagnosis codes. Moreover, the ACG system is built for population-level risk stratification, not precise individual-level prediction. Nevertheless, regression models solely based on ADGs have been developed before (23, 27, 41). Due to the availability of only overall accuracy metrics for discrimination and calibration, the potential for making direct comparisons to those previous studies is limited. For instance, one study (41) used ADGs for mortality prediction and had a c-statistic of 0.84, but with a 3.1% outcome event rate these metrics might not fully capture the model’s performance. Despite the limitations in model performance, we argue that our case study reflects the kinds of imperfect conditions found in real-world data upon which predictive models are often developed. Moreover, feature-level biases are largely independent of model performance and can emerge even when predictive accuracy is suboptimal. The use of SV remains valid for comparative purposes where relative differences can provide meaningful insights into potential sources of disparity in EHR data, even if the model’s overall predictive accuracy is limited. Any observed differences between the two datasets, such as with ADG 11 and ADG 32, reflect genuine differences in how the model utilizes features. Without such diagnostic analyses, fairness interventions risk being based on untested or overly simplistic assumptions about the dataset. Consequently, our FL-BIAS practice enables more effective (causal) mitigation strategies for algorithmic bias.

Yet, as argued by Corbett-Davies et al. (10), any technical results in the domain of algorithmic fairness must be made relevant to real-world contexts. A multidisciplinary team is needed to uncover underlying data-generating processes, such as variation in clinical recording practices, non-random missingness, or unmeasured variables affecting both health status and healthcare utilization. Moreover, the use of EHR for observational research is prone to various types of bias (42), which could distort interpretation. Stakeholder input is essential to determine whether observed differences reflect true disparities or spurious associations. Ultimately, algorithmic fairness assessments require a close interplay between in-depth data analysis and human interpretation. Therefore, our diagnostic framework operationalizes principles from Critical Data Studies, which emphasize the non-neutrality of data, by promoting reflexivity on data and facilitating sensemaking at the feature level, through which contextual awareness can emerge (43). We encourage future work to include additional analyses, for instance by testing different machine learning model architectures, comparing calibration curves as demonstrated by (44) or enhancing methodological rigor through bootstrapping, as applied by (21) for SV analyses.

## Conclusion

In-depth analyses of databases are key for uncovering the underlying causes of algorithmic bias in datasets and for promoting equity in healthcare delivery. This study introduced the Feature-Level Bias Identification And Sensemaking (FL-BIAS) diagnostic approach for detecting bias at the feature level. We demonstrated the use of FL-BIAS in a case study using Dutch primary care EHR data linked with Statistics Netherlands data, developing models based on ADGs from the Johns Hopkins ACG system. We observed modest disparities in overall model performance but notable differences in the relevance of specific ADG features across subgroups. For example, ADG 32 (Malignancy) had a stronger association with hospitalization among non-Western patients, potentially reflecting more advanced disease state.

These results highlight the need for deeper engagement with the data-generating process, including awareness of potential confounders such as cancer screening participation, when assessing algorithmic fairness. Such disparities might also be addressed through more granular feature engineering, such as splitting the feature for malignancy into severity-based categories (e.g., early-stage vs. advanced malignancy) to better capture clinical heterogeneity across populations. Healthcare organizations should implement routine fairness diagnostic approaches and involve multidisciplinary teams to contextualize algorithmic bias patterns and develop targeted interventions to reduce health disparities. FL-BIAS provides a practical framework for this essential work.

## Data Availability

The data supporting this study are subject to strict privacy regulations and are available from Statistics Netherlands (CBS) under specific conditions. Access to these microdata is possible for researchers who meet the criteria for confidential data access as set by CBS.

## Acknowledgments

We would like to thank Dr. Yara Bachour, MD, for her valuable clinical insights into the interpretation of ADG 32 and its relation to observed differences in cancer screening.

## Supporting information

**S1 Table. Model specifications. Contains detailed model parameters and hyperparameter grid search settings**.

**S2 Table. Overview prevalence ADG variables for Dutch and non-Western populations. Includes point prevalence differences and p-values from Pearson’s Chi-square Test**

**S3 File. Multiple Correspondence Analysis. Sample population characteristics for MCA analysis and Eigenvalues of the first ten dimensions.**

**S4 Table. Causal Mediation Analysis. Effect measures of the influence of the independent variable (migration background) on the dependent variable (Hospitalization) through the mediation variable (SES. Provides Average Causal Mediated Effect, Average Direct Effect and Rho Value.**

**S5 Table. Matthews correlation coefficients. Values for Dutch and non-Western logistic regression models, Rx-MG model, and XGBoost model.**

**S6 Table. Feature influence. Variance Inflation Factor (VIF) indicates the extent of multicollinearity in the dataset. Overview of coefficients of the logistic regression models with 32 ADGs, trained on the full population, Dutch population, and non-Western population.**

**S7 Fig. Correlation heatmap of ADG features.Visualization of correlations between ADG variables.**

**S8 Table. Kernel Shapley Values. overview global shapley values using Kernal SHAP to calculate feature contributions to deviations from a baseline prediction (“none”).**

**S9 Table. Overview global Cohort Shapley Values to calculate feature contributions to residuals and false negative cases**.

