## Supplementary material for "Refocusing Algorithmic Fairness on Feature-Level Bias: A Diagnostic Approach Using Dutch EHR Data": S1

### **S1. Model specifications**

| **Model** | **Training data** | **Feature transformation** | **Parameters** |
| --- | --- | --- | --- |
| *Logistic Regression - 32 ADGs* | 84.654 patients  (71.062 Dutch,  13.592 non-Western) | One-hot encoded variables: Age (binned), SES, Sex | L2 regression, class weight = balanced |
| *Logistic Regression - 32 ADGs Dutch* | 71.062 Dutch patients | One-hot encoded variables: Age (binned), SES, Sex | L2 regression, class weight = balanced |
| *Logistic Regression - 32 ADGs Non-Western* | 13.592 non-Western  patients | One-hot encoded variables: Age (binned), SES, Sex | L2 regression, class weight = balanced |
| *Logistic Regresion - major ADGs* | 84.654 patients (\  (71.062 Dutch,  13.592 non-Western) | One-hot encoded variables: Age (binned), SES, Sex | L2 regression, class weight = balanced |
| *Logistic regression- Rx-MG* | 84.654 patients  (71.062 Dutch,  13.592 non-Western) | One-hot encoded variables: Age (binned), SES, Sex | L2 regression, class weight = balanced |
| *XGBoost - 32 ADGs* | 84.654 patients  (71.062 Dutch,  13.592 non-western) | One-hot encoded variables: Age (binned), SES, Sex | nrounds = 300,  max_depth = 10  eta = 0.3  gamma = 0,  colsample_bytree = 0.9  min_child weight=1  subsample = 1  scale_pos_weight = 10  objective= binary:logistic  threshold = 0.5 |

*Overview of model specifications. XGBoost hyperparameters were optimized using grid search*
