## Supplementary material for "Refocusing Algorithmic Fairness on Feature-Level Bias: A Diagnostic Approach Using Dutch EHR Data": S2

### **S2. Prevalence ADG variables**

| ADG | Dutch  n (%) | Non-Western  n(%) | Percentage point difference | p-values from Pearson's Chi-dquare Test |
| --- | --- | --- | --- | --- |
| 1 Time Limited: Minor | 34254 (36.2) | 5483 (30.1) | 6.1% | <0.001 |
| 2 Time Limited: Minor-Primary Infections | 34902 (36.9) | 6708 (36.8) | 0.1% | 0.774 |
| 3 Time Limited: Major | 2743 ( 2.9) | 405 ( 2.2) | 0.7% | <0.001 |
| 4 Time Limited: Major-Primary Infections | 4108 ( 4.3) | 575 ( 3.2) | 1.1% | <0.001 |
| 5 Allergies | 13805 (14.6) | 3677 (20.2) | -5.6% | <0.001 |
| 6 Asthma | 6065 ( 6.4) | 1169 ( 6.4) | 0.0 % | 1.000 |
| 7 Likely to Recur: Discrete | 29148 (30.8) | 6541 (35.9) | -5.1% | <0.001 |
| 8 Likely to Recur: Discrete-Infections | 19866 (21.0) | 3286 (18.0) | -3.0% | <0.001 |
| 9 Likely to Recur: Progressive | 2883 ( 3.0) | 459 ( 2.5) | 0.5% | <0.001 |
| 10 Chronic Medical: Stable | 35010 (37.0) | 6337 (34.7) | 2.3% | <0.001 |
| 11 Chronic Medical: Unstable | 13831 (14.6) | 2125 (11.6) | 3.0% | <0.001 |
| 12 Chronic Specialty: Stable-Orthopedic | 4812 ( 5.1) | 951 ( 5.2) | -0.1% | 0.485 |
| 13 Chronic Specialty: Stable-Ear, Nose, Throat | 3075 ( 3.2) | 404 ( 2.2) | 1.0% | <0.001 |
| 14 Chronic Specialty: Stable-Eye | 5841 ( 6.2) | 1030 ( 5.6) | 0.6% | 0.007 |
| 16 Chronic Specialty: UnstableOrthopedic | 5244 ( 5.5) | 954 ( 5.2) | 0.3% | 0.093 |
| 17 Chronic Specialty: Unstable-Ear, Nose, Throat | 396 ( 0.4) | 42 ( 0.2) | 0.2% | <0.001 |
| 18 Chronic Specialty: Unstable-Eye | 1455 ( 1.5) | 235 ( 1.3) | 0.2% | 0.012 |
| 20 Dermatologic | 29782 (31.5) | 4874 (26.7) | 4.8% | <0.001 |
| 21 Injuries/Adverse Effects: Minor | 26756 (28.3) | 4637 (25.4) | 2.9% | <0.001 |
| 22 Injuries/Adverse Effects: Major | 13377 (14.1) | 1911 (10.5) | 3.6% | <0.001 |
| 23 Psychosocial: Time Limited, Minor | 14982 (15.8) | 2661 (14.6) | 1.2% | <0.001 |
| 24 Psychosocial: Recurrent or Persistent, Stable | 18546 (19.6) | 3552 (19.5) | 0.1% | 0.694 |
| 25 Psychosocial: Recurrent or Persistent, Unstable | 2350 ( 2.5) | 489 ( 2.7) | -0.2% | 0.126 |
| 26 Signs/Symptoms: Minor | 64509 (68.2) | 12756 (69.9) | -1.7% | <0.001 |
| 27 Signs/Symptoms: Uncertain | 62493 (66.0) | 12183 (66.8) | -0.8% | 0.055 |
| 28 Signs/Symptoms: Major | 17104 (18.1) | 3326 (18.2) | -0.1% | 0.624 |
| 29 Discretionary | 15142 (16.0) | 2232 (12.2) | 3.8% | <0.001 |
| 30 See and Reassure | 12406 (13.1) | 2660 (14.6) | -1.5% | <0.001 |
| 31 Prevention/  Administrative | 33368 (35.3) | 6210 (34.0) | 1.3% | 0.002 |
| 32 Malignancy | 5294 ( 5.6) | 477 ( 2.6) | 3.0% | <0.001 |

*Overview Exploratory Data Analysis ADG. The prevalence of ADGs is presented for both the Dutch and non-Western populations, with the p-value indicating the level of statistical significance in the differences observed.*
