## Supplementary material for "Refocusing Algorithmic Fairness on Feature-Level Bias: A Diagnostic Approach Using Dutch EHR Data": S3

### **S3. Multiple Correspondence Analysis**

|  | Dutch | non-Western |
| --- | --- | --- |
| n | 6681 | 6681 |
| Age (mean (SD)) | 45.81 (19.92) | 44.06 (18.09) |
| GP visits (mean (SD)) | 3.53 (4.37) | 3.60 (4.13) |
| Male (%) | 3236 (48.4) | 3127 (46.8) |
| SES (%) | | |
| High | 2227 (33.3) | 2227 (33.3) |
| Low | 2227 (33.3) | 2227 (33.3) |
| Middle | 2227 (33.3) | 2227 (33.3) |
| Hospitalized (%) | 645 ( 9.7) | 645 ( 9.7) |

*Sample population characteristics for MCA analysis*

|  | Eigenvalues Dutch | Eigenvalues  non-Western |
| --- | --- | --- |
| Dim.1 | 0.105 | 0.094 |
| Dim.2 | 0.049 | 0.047 |
| Dim.3 | 0.037 | 0.036 |
| Dim.4 | 0.036 | 0.035 |
| Dim.5 | 0.034 | 0.034 |
| Dim.6 | 0.033 | 0.033 |
| Dim.7 | 0.033 | 0.032 |
| Dim.8 | 0.032 | 0.032 |
| Dim.9 | 0.031 | 0.031 |
| Dim.10 | 0.031 | 0.031 |

*Eigenvalues of the first ten dimensions obtained from the Multiple Correspondence Analysis (MCA) for Dutch and non-Western populations. Similar eigenvalue distributions indicate comparable underlying data structures between the two groups.*
