## Supplementary material for "Refocusing Algorithmic Fairness on Feature-Level Bias: A Diagnostic Approach Using Dutch EHR Data": S4

### **S4. Causal Mediation Analysis**

| Mediation  analysis | Estimate | 95% CI lower | 95% CI upper | p-value |
| --- | --- | --- | --- | --- |
| ACME (control) | 0.005758 | 0.004824 | 0.01 | <2e-16 |
| ACME (treated) | 0.005695 | 0.004806 | 0.01 | <2e-16 |
| ACME Sensitivity analysis Rho value | - 0.1 |  |  |  |
| ADE (control) | -0.001442 | -0.005332 | 0.00 | 0.68 |
| ADE (treated) | -0.001505 | -0.005575 | 0.00 | 0.68 |
| Proportion mediated | 1.346575 | 0.529911 | 9.13 | 0.04 |

*Results Mediation Analysis with R library Mediation based on 100 bootstrap simulations. ACME: Average Causal Mediated Effect* *measures influence of the independent variable (migration background) on the dependent variable (Hospitalization) through mediation variable (SES). ADE: Average Direct Effect measures direct influence of migration background on hospitalization. The Rho value indicates the extent of pre-treatment mediator-outcome confounding required to yield a null ACME. The higher the value of Rho (within the range of -1 to 1), the more robust the ACME estimate.*
