## Supplementary material for "Refocusing Algorithmic Fairness on Feature-Level Bias: A Diagnostic Approach Using Dutch EHR Data": S5

### **S5. Matthews Correlation coefficients**

|  | Dutch (23566) | Non western (4652) |
| --- | --- | --- |
| MCC LR-32 ADGs | 0.214 | 0.225 |
| *MCC Parity* | *1* | *1,051* |
| MCC LR Rx-MG | 0.236 | 0.222 |
| *MCC Parity* | *1* | *0.941* |
| MCC XGBoost | 0.170 | 0.185 |
| *MCC Parity* | *1* | *1,090* |

*Matthews Correlation Coefficient (MCC)*
