## Supplementary material for "Refocusing Algorithmic Fairness on Feature-Level Bias: A Diagnostic Approach Using Dutch EHR Data": S6

### **S6. Feature Influence**

| Feature | VIF | LR -32 coeffs total population | LR-32 coeffs Dutch | LR -32 coeffs non-Western |
| --- | --- | --- | --- | --- |
| Age 12-34 | 4.720* | -0.613 | -0.628 | -0.416 |
| Age 35-54 | 5.172* | -0.358 | -0.359 | -0.208 |
| Age 55-69 | 5.069* | 0.109 | 0.115 | 0.205 |
| Age 70-79 | 3.015* | 0.302 | 0.305 | 0.415 |
| Age 80+ | 1.153* | 0.044 | 0.053 | 0.004 |
| Female | 1.430* | -0.254 | -0.249 | -0.021 |
| Male | 1.397* | -0.263 | -0.265 | 0.021 |
| High SES | 4.720* | -0.248 | -0.258 | -0.022 |
| Middle SES | 5.172* | -0.145 | -0.154 | 0.049 |
| Low SES | 5.069* | -0.124 | -0.102 | -0.027 |
| 1 Time Limited: Minor | 1.112 | -0.029 | -0.058 | 0.129 |
| 2 Time Limited: Minor-Primary Infections | 1.130 | 0.104 | 0.092 | 0.170 |
| 3 Time Limited: Major | 1.017 | 0.319 | 0.306 | 0.423 |
| 4 Time Limited: Major-Primary Infections | 1.025 | -0.064 | -0.072 | -0.047 |
| 5 Allergies | 1.060 | 0.057 | 0.052 | 0.087 |
| 6 Asthma | 1.032 | 0.135 | 0.168 | -0.033 |
| 7 Likely to Recur: Discrete | 1.145 | 0.110 | 0.130 | 0.006 |
| 8 Likely to Recur: Discrete-Infections | 1.179 | 0.117 | 0.113 | 0.156 |
| 9 Likely to Recur: Progressive | 1.075 | 0.550 | 0.537 | 0.568 |
| 10 Chronic Medical: Stable | 1.358 | 0.372 | 0.364 | 0.427 |
| 11 Chronic Medical: Unstable | 1.241 | 0.518 | 0.520 | 0.515 |
| 12 Chronic Specialty: Stable-Orthopedic | 1.031 | 0.155 | 0.180 | 0.021 |
| 13 Chronic Specialty: Stable-Ear, Nose, Throat | 1.042 | 0.105 | 0.120 | 0.042 |
| 14 Chronic Specialty: Stable-Eye | 1.156 | 0.150 | 0.160 | 0.097 |
| 16 Chronic Specialty: UnstableOrthopedic | 1.034 | 0.235 | 0.231 | 0.238 |
| 17 Chronic Specialty: Unstable-Ear, Nose, Throat | 1.005 | 0.368 | 0.458 | -0.571 |
| 18 Chronic Specialty: Unstable-Eye | 1.018 | 0.045 | -0.022 | 0.431 |
| 20 Dermatologic | 1.084 | -0.023 | -0.017 | -0.053 |
| 21 Injuries/Adverse Effects: Minor | 1.081 | 0.130 | 0.155 | -0.003 |
| 22 Injuries/Adverse Effects: Major | 1.046 | 0.135 | 0.133 | 0.162 |
| 23 Psychosocial: Time Limited, Minor | 1.074 | 0.049 | 0.050 | 0.044 |
| 24 Psychosocial: Recurrent or Persistent, Stable | 1.091 | 0.043 | 0.049 | 0.009 |
| 25 Psychosocial: Recurrent or Persistent, Unstable | 1.028 | 0.093 | 0.093 | 0.085 |
| 26 Signs/Symptoms: Minor | 1.149 | 0.259 | 0.272 | 0.196 |
| 27 Signs/Symptoms: Uncertain | 1.168 | 0.301 | 0.301 | 0.304 |
| 28 Signs/Symptoms: Major | 1.107 | 0.264 | 0.270 | 0.226 |
| 29 Discretionary | 1.079 | 0.184 | 0.152 | 0.396 |
| 30 See and Reassure | 1.063 | 0.073 | 0.054 | 0.180 |
| 31 Prevention/  Administrative | 1.128 | -0.041 | -0.049 | 0.007 |
| 32 Malignancy | 1.089 | 0.530 | 0.510 | 0.742 |
| 33 Pregnancy | 1.112 | 0.716 | 0.647 | 0.970 |
| 34 Dental | 1.130 | 0.125 | 0.074 | 0.339 |

*Variance Inflation Factor (VIF) column indicates the extent of multicollinearity in the entire database.Overview of the coefficients of the Logistic Regression models with 32 ADGs, trained on full population (84.654 patients), Dutch population (71.062 patients) and non-Western population (13.592 patients). *High VIFs for categorical variables (e.g., Age, SES) are expected due to inclusion of all dummy variables; in practice, one category would be omitted (reference category) to avoid multicollinearity.*
