## Supplementary material for "Refocusing Algorithmic Fairness on Feature-Level Bias: A Diagnostic Approach Using Dutch EHR Data": S7

### **S8. Kernel Shapley Values**

| Kernel Shap | Dutch* | | | Non- Western* | | |
| --- | --- | --- | --- | --- | --- | --- |
|  | Means | St deviation | Sum absolute | Means | St deviation | Sum absolute |
| none | 0.5 | 0 | 1000 | 0.5 | 0 | 1000 |
| Predicted | 0.5069 | 0.182 | 1013.71 | 0.4706 | 0.158 | 941.26 |
| Age | 0.0007 | 0.101 | 171.25 | -0.0027 | 0.089 | 146.23 |
| Sex | 0.0006 | 0.051 | 102.02 | -0.0027 | 0.051 | 101.55 |
| SES | 0.0007 | 0.022 | 39.40 | -0.0026 | 0.016 | 27.51 |
| 3 Time Limited: Major | 0.0006 | 0.034 | 26.31 | -0.0027 | 0.025 | 21.14 |
| 4 Time Limited: Major-Primary Infections | 0.0006 | 0.006 | 3.94 | -0.0027 | 0.008 | 11.08 |
| 9 Likely to Recur: Progressive | 0.0006 | 0.042 | 34.58 | -0.0027 | 0.040 | 36.63 |
| 11 Chronic Medical: Unstable | 0.0004 | 0.092 | 148.92 | -0.0027 | 0.082 | 124.60 |
| 16 Chronic Specialty: UnstableOrthopedic | 0.0006 | 0.029 | 28.54 | -0.0027 | 0.031 | 35.24 |
| 22 Injuries/Adverse Effects: Major | 0.0006 | 0.022 | 30.90 | -0.0027 | 0.020 | 30.21 |
| 25 Psychosocial: Recurrent or Persistent, Unstable | 0.0006 | 0.010 | 5.41 | -0.0027 | 0.011 | 12.45 |
| 32 Malignancy | 0.0007 | 0.054 | 57.31 | -0.0027 | 0.040 | 37.61 |

(*)Subset of data used, with 1000 positive event (hospitalization) and 1000 negative event (no hospitalization)

*Overview global Shapley Values using Kernal Shapley Vlaues to calculate responsiveness of features to deviations from a baseline prediction (“none”)*
