## Supplementary material for "Refocusing Algorithmic Fairness on Feature-Level Bias: A Diagnostic Approach Using Dutch EHR Data": S8

### **S9. Cohort Shapley Values**

|  | Residuals (n=27184) | | | False Negatives (subset with hospitalization outcome) ** | | |
| --- | --- | --- | --- | --- | --- | --- |
| **Cohort global shapley values -absolute sum** | Full | 50% Dutch | 50%  Non-  western | Full | 50% Dutch | 50%  Non- western |
| Age | 1179.41 | 619.09 | 560.32 | 596.82 | 286.92 | 309.90 |
| Sex | 746.55 | 387.34 | 359.21 | 149.75 | 68.62 | 81.13 |
| SES | 222.45 | 105.92 | 116.53 | 59.22 | 31.65 | 27.57 |
| 3 Time Limited: Major | 123.43 | 66.09 | 57.34 | 48.86 | 24.35 | 24.51 |
| 4 Time Limited: Major-Primary Infections | 101.43 | 56.41 | 45.02 | 21.3 | 10.58 | 10.72 |
| 9 Likely to Recur: Progressive | 135.73 | 71.54 | 64.20 | 88.6 | 41.45 | 47.15 |
| 11 Chronic Medical: Unstable | 617.25 | 325.86 | 291.39 | 457.72 | 216.65 | 241.07 |
| 16 Chronic Specialty: UnstableOrthopedic | 240.36 | 122.43 | 117.93 | 121.93 | 55.39 | 66.54 |
| 22 Injuries/Adverse Effects: Major | 300.23 | 158.89 | 141.34 | 122.26 | 59.70 | 62.56 |
| 25 Psychosocial: Recurrent or Persistent, Unstable | 72.11 | 36.99 | 35.12 | 13.27 | 6.26 | 7.01 |
| 32 Malignancy | 221.70 | 133.30 | 88.40 | 169.32 | 94.54 | 74.78 |

(**)Subset of dataset with only hospitalization outcome resulted into 2604 individuals. Balance was slightly skewed (1359 Dutch and 1235 Non-Western), so equal groups of 1235 were sampled.

*Overview global Cohort Shapley Values to calculate feature contributions to residuals and false negative cases*
